# Modelling the potential spread of Clade Ib MPXV in an Asia Pacific city

**DOI:** 10.1101/2024.09.10.24313384

**Authors:** Shihui Jin, Gregory Gan, Akira Endo, Kiesha Prem, Rayner Kay Jin Tan, Jue Tao Lim, Keisuke Ejima, Borame L Dickens

## Abstract

**Background:** The ongoing 2024 mpox outbreak in several African countries, driven by the novel Clade 1b strain, has resulted in imported cases being reported in Sweden and Thailand. The Asia Pacific (APAC) region, with little exposure to previous mpox waves, is particularly vulnerable to local transmission triggered by importation. While this highlights the importance of early preparedness, current knowledge of the virus’s transmission dynamics remains too limited to effectively inform policy-making and resource planning.

**Methods:** A compartmental model was constructed to characterise the potential mpox transmission triggered by importation in an APAC city. Outbreaks were simulated under diverse hypothetical scenarios considering transmission mechanisms, different affected subpopulations, levels of disease transmissibility, and importation frequencies. The impacts of various non-pharmaceutical interventions (NPIs) including isolation, and vaccination strategies were projected and compared.

**Findings:** Up to 30% of the population would be infected in the scenario of high sexual and moderate non-sexual transmissibility in the general community, with minimal impact from importation frequency on outbreak size and healthcare burden. The sequential introduction of mandatory home isolation for diagnosed cases, pre-departure screening of international arrivals, and contact tracing were projected to lower the peak outbreak size by 35%, 0.04%, and 1.1%, respectively, while the reduction would be 30% with proper vaccination by prioritising the sexually active group. These measures would significantly decrease disease morbidity and mortality rates, thereby alleviating the disease’s pressure on healthcare systems.

**Interpretation:** The potential mpox outbreak in the APAC setting could be alleviated through strong surveillance and timely response from stakeholders. NPIs are recommended over vaccination for outbreak management due to their demonstrated effectiveness and practicability.

## 1. Introduction

An outbreak caused by Clade I monkeypox virus (MPXV) was estimated to have emerged in September 2023 in South Kivu province, Democratic Republic of the Congo (DRC), with transmission likely driven primarily by sexual contact. The novel strain of the outbreak, later designated as Clade Ib, resulted in surging cases reported in the DRC, alongside the endemic Clade Ia already circulating in other provinces, and quickly spread to neighbouring African countries, including Burundi, Rwanda, Uganda, and Kenya. In response to the rapid spread of Clade Ib in Africa, the World Health Organisation re-declared mpox as a public health emergency of international concern (PHEIC), aiming to raise the global awareness and prevent a repeat of the 2022 mpox outbreak, whose global circulation was mainly the consequence of initial negligence in surveillance and prevention [1].

During the 2022 wave caused by Clade IIb MPXV, over 90% of the cases reported were from Europe and the Americas. In contrast, the Asia Pacific (APAC) region, including the Western Pacific and Southeast Asia, reported fewer than 5% of all Clade IIb cases [2]. While possibility of high under-ascertainment exists due to the limited surveillance efforts and reluctance in seeking medical help due to stigmas associated with the infection [3], it is evident that the population with active immunity against the virus should be much smaller in APAC compared to other regions of the world with higher cumulative prevalence and/or better vaccine supply. This can make the region extremely vulnerable to potential Clade Ib mpox outbreaks once the virus begins to circulate globally.

While Clade IIb MPXV was predominantly spread through sexual transmission within the gay, bisexual and other men-who-have-sex-with-men (GBMSM) community [4], evidence has shown that Clade Ib, the strain driving the ongoing 2024 mpox outbreak in Africa, can be transmitted through non-sexual close contacts apart from the well-documented sexual means [5,6]. This raises the possibility that a future mpox outbreak in APAC could affect the general population, with non-sexual transmission routes suspected to be more prevalent compared to the previous 2022 outbreak.

Throughout the COVID-19 pandemic, territories in the APAC region gained valuable experience in implementing non-pharmaceutical interventions (NPIs) [7,8]. Social distancing and border control measures, which limited travel and reduced interpersonal contact, effectively contained the disease transmission [9]. These successful stories offer hope for the authorities in this area to leverage NPIs to curb a potential mpox outbreak, should it be widespread in the general community. Furthermore, the absence of firm evidence for respiratory transmission [10] for MPXV implies that effective outbreak management might be achieved through less stringent NPIs, such as isolating infected individuals, compared to those implemented during the COVID-19 pandemic.

Nevertheless, few NPIs were applied to manage the 2022 mpox outbreak, which affected only a limited subpopulation. This leaves us great uncertainty about their true effectiveness and sufficiency in averting a more widespread mpox outbreak once it establishes itself in the general population. Furthermore, the lack of comprehensive surveillance data have also brought great unsureness surrounding the possible transmission patterns of Clade Ib MPXV, especially in the APAC setting without history of mpox outbreaks.

To address these gaps, we performed this study to model various potential mpox outbreak scenarios in Singapore, a city-state in Southeast Asia closely connected to the international community. We hypothesised different subpopulations as the primary drivers and vulnerable groups of the importation-triggered outbreak and simulated epidemic trajectories utilising a modified deterministic susceptible-exposed-infectious-recovered (SEIR) model. We further projected the impacts of NPIs and vaccination on curbing the outbreak and reducing the disease burden. These results collectively provide insights into factors shaping the outbreak, informing policy-makers of strategies to pre-empt or mitigate mpox transmission in Singapore and other territories in the APAC region.

## 2. Methods

### 2.1 SEIR-style transmission model

We adapted a deterministic SEIR compartmental model to characterise potential disease transmission patterns, as shown in Figure 1. Additional compartments, including Quarantined (*Q*), Diagnosed (*C*), Hospitalised (*H*), Admitted to ICU (*U*), and Dead (*D*), were incorporated into the model to facilitate a more accurate reflection of a real-world outbreak, with disease surveillance, variations in disease burden among individuals, and the probable presence of intervention measures.

**Figure 1.**
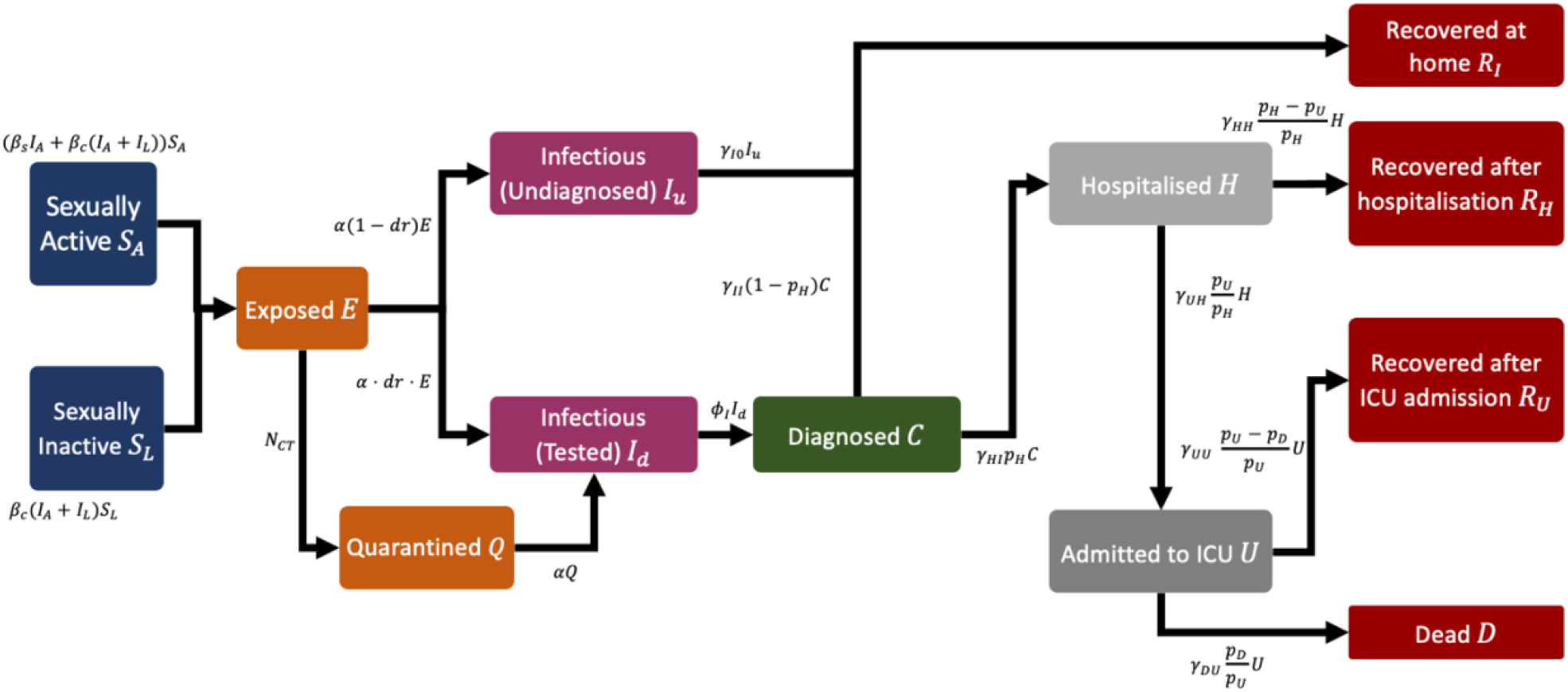
Model schematic. *I*_*A*_ = *I*_*uA*_ + (1 − *c*_*iso*_)(*I*_*dA*_ + *C*_*A*_) and *I*_*L*_ = *I*_*uL*_ + (1 − *c*_*iso*_)(*I*_*dL*_ + *C*_*L*_) refer to the effective infectious sexually active and inactive subpopulation, respectively, where *c*_*iso*_ is the reduced capability of disease spreading due to quarantine or isolation and equals 1 in the absence of them. *N*_*CT*_ is the number of traced close contacts, whose value depends on contact tracing capacity, number of newly diagnosed cases and existing exposed population. See Table S1 for more details for these parameters.

Due to the disparity in transmission potential of mpox via sexual and non-sexual activities, we segregated the affected population into two cohorts: sexually active (*A*) and inactive (*L*) individuals, assuming the latter could only contract or spread the infection through non-sexual means. Diagnostic rate and disease severity were not distinguished between these two groups.

Infections who underwent clinical testing would be diagnosed and reported, while those untested were assumed to have mild symptoms and recover at home. Among the diagnosed cases, some would also recover at home, while others would be hospitalised as the disease progressed. Those with severe symptoms would be admitted to intensive care units (ICU) after hospitalisation, where they would be either overcome or succumb to mpox. We also assumed that deaths occurred only among ICU admissions.

Intervention measures built into the model include isolating diagnosed cases and quarantining infections who were tested but yet diagnosed as well as the close contacts of confirmed cases. Both groups were presumed to have limited daily interactions with others. Please refer to section 2.4 and the Supplementary Information for more details.

### 2.2 Scenarios for outbreak simulations

Utilising the established compartmental model, we simulated potential mpox outbreaks under a total of five hypothetical scenarios due to the limited knowledge we had of the strain in circulation at the time of this study. These scenarios fell into two categories: transmission limited to specific high-risk subpopulations and their close contacts (Scenario 1 and 2) and transmission in the general population (Scenario 3a–c). They include:

1. Outbreak driven by GBMSM community in Singapore,
2. Outbreak driven by sex workers in Singapore,
3. Outbreak in general population, with
  a. limited local transmission,
  b. moderate local transmission,
  c. widespread local transmission.

Scenario 1, 2, and 3 primarily differed in the sizes of the population affected by the outbreak, and its composition of sexually active (A) and inactive (L) subpopulations, while the two scenarios for transmission in the general population (i.e., Scenario 3a–c) had identical population sizes but varied in their parameter values for disease transmissibility in the general community (*R*_*c*_). Individuals vaccinated against small pox, particularly those aged over 45 years, were considered immune to the outbreak due to the high efficacy of the vaccine and were therefore excluded from the affected population [11,12]. However, we did not account for cross protection from exposure to previous mpox outbreaks, as the area was barely affected by those waves.[2] Please refer to Table S3 for specific values of the model parameters for individual scenarios.

It should be noted that we assumed a 50% reduction in contacts between the susceptible population and infections who sought medical assistance. In a sensitivity analysis, we explored how variations in this level of spontaneous isolation would shape the outbreak. Nonetheless, no NPIs (e.g., mandatory isolation, quarantine, or contact tracing) were implemented in any of these five scenarios. Furthermore, we allowed for importation through international arrivals, assuming five imported infections every three days. We also presumed the diagnostic rate of imported cases to be much lower, at half the level of that for the local population (i.e., 60%), to account for the potential reduced propensity of international travellers in seeking healthcare services [13]. However, this reduction in diagnostic rate would not inflate the ratios between diagnosed cases and hospitalisations, ICU admissions, or deaths among imported infections.

Scenario 3b, which we believe to be the most plausible representation of a future outbreak, was selected as the baseline scenario for predicting the impacts of disease transmission potential, importation volume, or diverse intervention strategies on morbidity and mortality. Its balanced and generic framework would allow for result extrapolation should other scenarios better characterise the future outbreak, making it the optimal candidate for modelling various potential outcomes. Henceforth, this scenario is referred to as the ‘baseline scenario’.

### 2.3 Disease transmissibility and importation influence assessment

Transmissibility of mpox was quantified by *R*_*s*_ and *R*_*c*_, representing the average number of secondary infections generated by an infector through sexual and non-sexual routes (e.g., close, personal contact), respectively, in a population with no previous exposure to the disease [14]. Due to the limited data available at the time of this study, it was challenging to pinpoint specific values for these two parameters. Therefore, we adopted a comprehensive approach, exploring a wide range of candidate values. With the belief that Clade Ib MPXV was primarily transmitted through sexual contact [15], we assumed *R*_*s*_ greater than *R*_*c*_ in most of the scenarios. Particularly, we let *R*_*s*_ to be one of 1.0, 2.5, 4.0, or 5.5, and *R*_*c*_ to be one of 0.3, 0.6, 0.9, or 1.2. These grid points were chosen for clarity and ease of presentation, but in a sensitivity analysis we further examined the impacts of *R*_*s*_ and *R*_*c*_ pairs on epidemic trajectories using values randomly sampled from continuous distributions (Figure S3).

To investigate how importation would shape local transmission dynamics, we considered four frequencies of importation events, ranked from low to high as follows:

I1. Monthly importation,

I2. Weekly importation, at the start of each week,

I3. Importation every three days,

I4. Daily importation.

For all the importation events, we assumed the number of imported cases per time to be five. All possible combinations of *R*_*s*_ and *R*_*c*_ candidate values were utilised to assess their collective impacts on the epidemic trajectory. The rest of the model parameters followed those in the baseline scenario.

### 2.4 Intervention effect projection

We evaluated two community-level NPIs: mandatory stay-at-home requirement for tested or diagnosed cases (henceforth referred to as ‘isolation’) and quarantine of their close contacts captured by contact tracing (henceforth referred to as ‘contact tracing’). These measures aimed at segregating the susceptible population from the detected infections and individuals exposed but not yet infectious, thereby greatly reducing the risk of transmission.

Nevertheless, we only assumed an additional 50% decrease in contacts due to these two NPIs compared to the aforementioned self-isolation, to account for potential within-household transmission (Table S4). Meanwhile, for border control, we proposed pre-departure screening for arrivals from high-risk countries, allowing entry exclusively to those with negative test results (henceforth referred to as ‘screening’). We assumed a 60% sensitivity for this pre-departure testing, but tested this assumption in a subsequent sensitivity analysis. Further details regarding these NPIs are elaborated in the Supplementary Information.

Given that these NPIs are frequently implemented alongside each other in real-world practices, we proposed three intervention strategies with distinct combinations of individual measures, ordered by their level of intensity:

C1. Isolation alone,

C2. Isolation and screening,

C3. Isolation, screening, and contact tracing.

Outbreaks during which these NPI combinations were in place were simulated using the same parameters as the baseline scenario, which served as the benchmark for comparison and effect evaluation. Additional sensitivity analyses were conducted for scenarios with extremely low or high risks of community spread.

We further investigated three different vaccine allocation plans to assess how effectively vaccination campaigns before the outbreak would mitigate disease outbreaks. Specifically, we fixed the increase in vaccine coverage of the total population at 5%, but varied the allocation of vaccines between the sexually active and inactive groups. The proposed plans are listed as below, and Table S5 in the Supplementary Information specified the vaccine uptake rates among the susceptible population in the two groups for each approach:

V1. All the vaccines administered by individuals belonging to the sexually active group,

V2. Equal vaccine uptake rate in both groups,

V3. Vaccine uptake higher in the sexually active group, with 75% of all doses administered to this group

We assumed the vaccine efficacy, *ve*, to be 66% [16]. Together with the vaccine coverage, *vacov*, of a specific population, this implied a 100(1 − *ve*)*vacov*% reduction in the susceptible population for the targeted group(s). To measure the effectiveness of these vaccination plans in reducing disease burden, we simulated outbreaks using various combinations of *R*_*s*_ and *R*_*c*_ values, as detailed in section 2.3. All the other parameters for simulation followed those in the baseline scenario. The model outputs were compared with those of the model with no vaccine rollout.

For all the aforementioned we simulated outbreaks over an 18-month (i.e., 540-day) period. We started each outbreak with none of the affected population exposed to or infected with Clade 1b MPVX, to mimic the current landscape in Singapore, where a potential outbreak would potentially be triggered by importation. Statistics to reflect disease burden, such as peak number of active hospitalised cases and total numbers of deaths, were summarised from the simulated trajectories. All the analyses and visualisation were performed using the R software [17].

## 3. Results

### 3.1 Hypothetical mpox outbreaks without interventions implemented

Large-scale mpox outbreaks were observed across all the five hypothetical scenarios in which no intervention strategies were applied. By the end of the 18 months following the initial importation event, proportions of infections in the affected population would be 78%, 92%, 41%, 46%, and 53% respectively.

The two scenarios (Scenarios 1 and 2) involving only a subpopulation shared similar outbreak sizes, with the maximum new infections being around 8,000. The burden on healthcare systems were also comparable. At its peak, the numbers of in-patient mpox infections staying in normal wards and ICU were expected to be as high as 8,500 and 4,000, respectively. These figures would at least triple when the general community was affected by the outbreak (Scenario 3a–c) and were positively correlated with the transmissibility of non-sexual transmission. For example, in the scenario with moderate disease transmission (Scenario 3b), up to 21,000 patients were infected in one day and nearly 27,000 hospital beds were occupied by patients with mpox (Figure 2).

**Figure 2.**
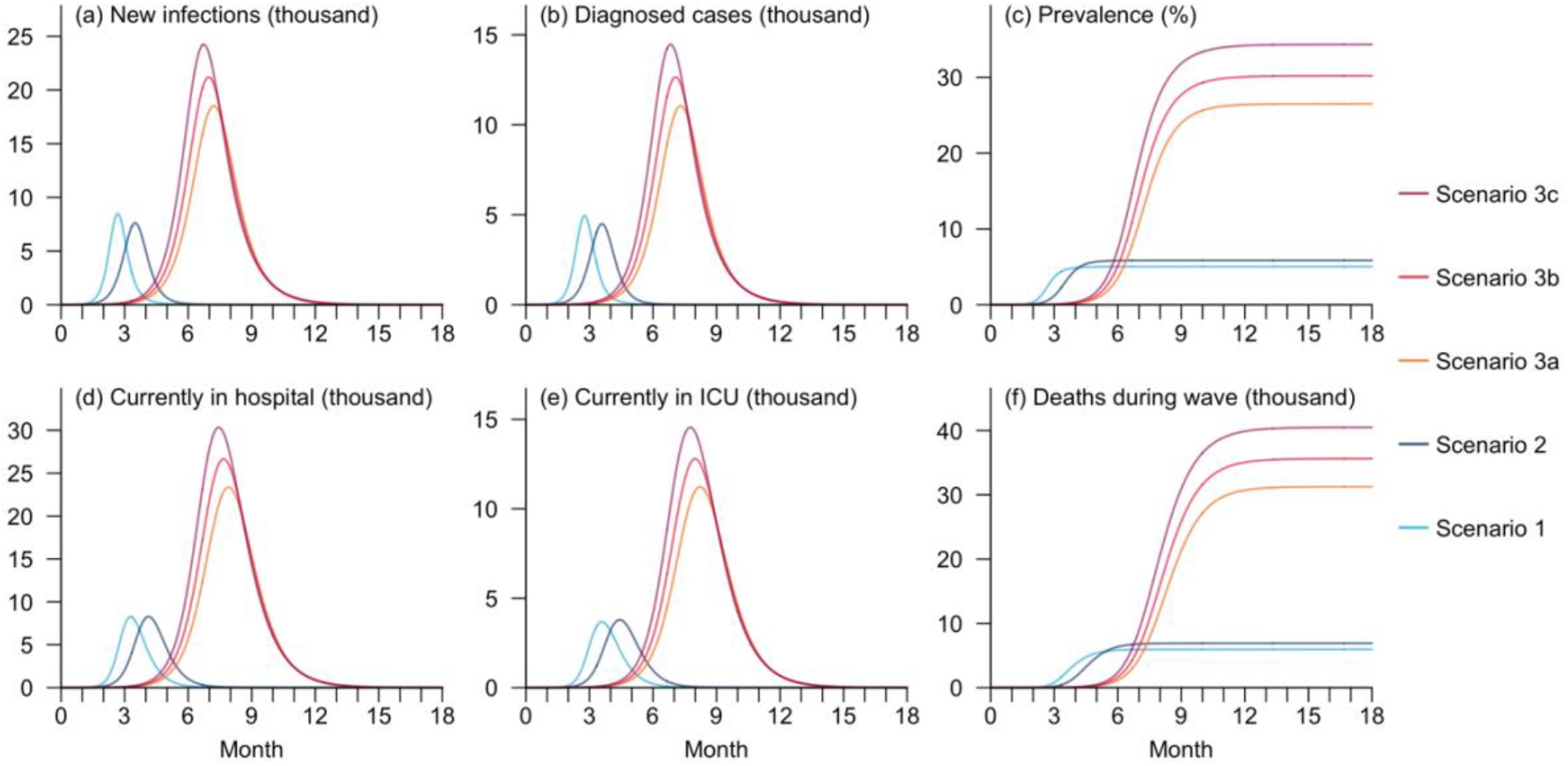
Disease burden and healthcare resource utilisation (HCRU) during mpox outbreaks in the five hypothetical scenarios. The indicators include number of new local infections (a) and diagnosed cases (b) on each day, disease prevalence in the overall local population (c), number of hospitalized cases in normal wards (d) and ICU occupancy (e), as well as cumulative number of deaths (f) over the 18 months following the initial importation event.

In each scenario, the outbreak peaked within six months and ended within one year from the initial importation, despite great variation in the timing. For epidemics driven by the GBMSM community (Scenario 1) or sex workers (Scenario 2), infections were projected to peak within a quarter due to the presumed high transmissibility of sexual transmission, while the wave’s peak would not occur until the fifth month for the transmission among the general population (Scenario 3a–c), with a maximum delay of one month due to reduced transmissibility. Furthermore, the delay between the peak of the infection trajectory and the highest healthcare resource utilisation (HCRU) was approximately 3 weeks across all the hypothetical scenarios (Figure 2).

### 3.2 Impacts of disease transmissibility and importation on simulated outbreaks

Variations in disease transmissibility would result in prominent changes in the outbreak scales. When *R*_*s*_, the metric for disease transmissibility through sexual contact, was 1.0, 2.5, and 5.5, the overall infection size at the end of 18 months would respectively be 0.04%, 41%, and 148% that of the baseline scenario where *R*_*s*_ = 4.0, assuming all other factors remained the same. Meanwhile, adjusting *R*_*c*_, the metric for disease transmissibility through non-sexual activities to 0.3, 0.9, and 1.2, would lead to infection sizes being 72%, 114%, and 130% that of the baseline scenario with *R*_*c*_ = 0.6. The trends were similar across diverse outbreak size indicators (Table S6).

Importation also played a role in shaping the local outbreaks. For instance, when *R*_*s*_ = 1.0 and *R*_*c*_ = 0.9, and the importation frequency was adjusted to monthly, weekly, and daily, the corresponding sero-prevalence in the general population by 18 months from the initial importation was 90%, 98%, and 102% of that in the scenario with an importation frequency of once per three days, respectively (Table S7). However, the impact of importation frequency was hardly observable in scenarios with relatively low transmissibility of sexual transmission (i.e., *R*_*s*_). In cases where *R*_*s*_ was sufficiently high, the transmission was quickly dominated by local infections, minimising the influence of the imported infections with an assumed size of five per event in our simulations and thus making the differences between scenarios with diverse importation frequencies hardly distinguishable (Figure 3, Figure S1).

**Figure 3.**
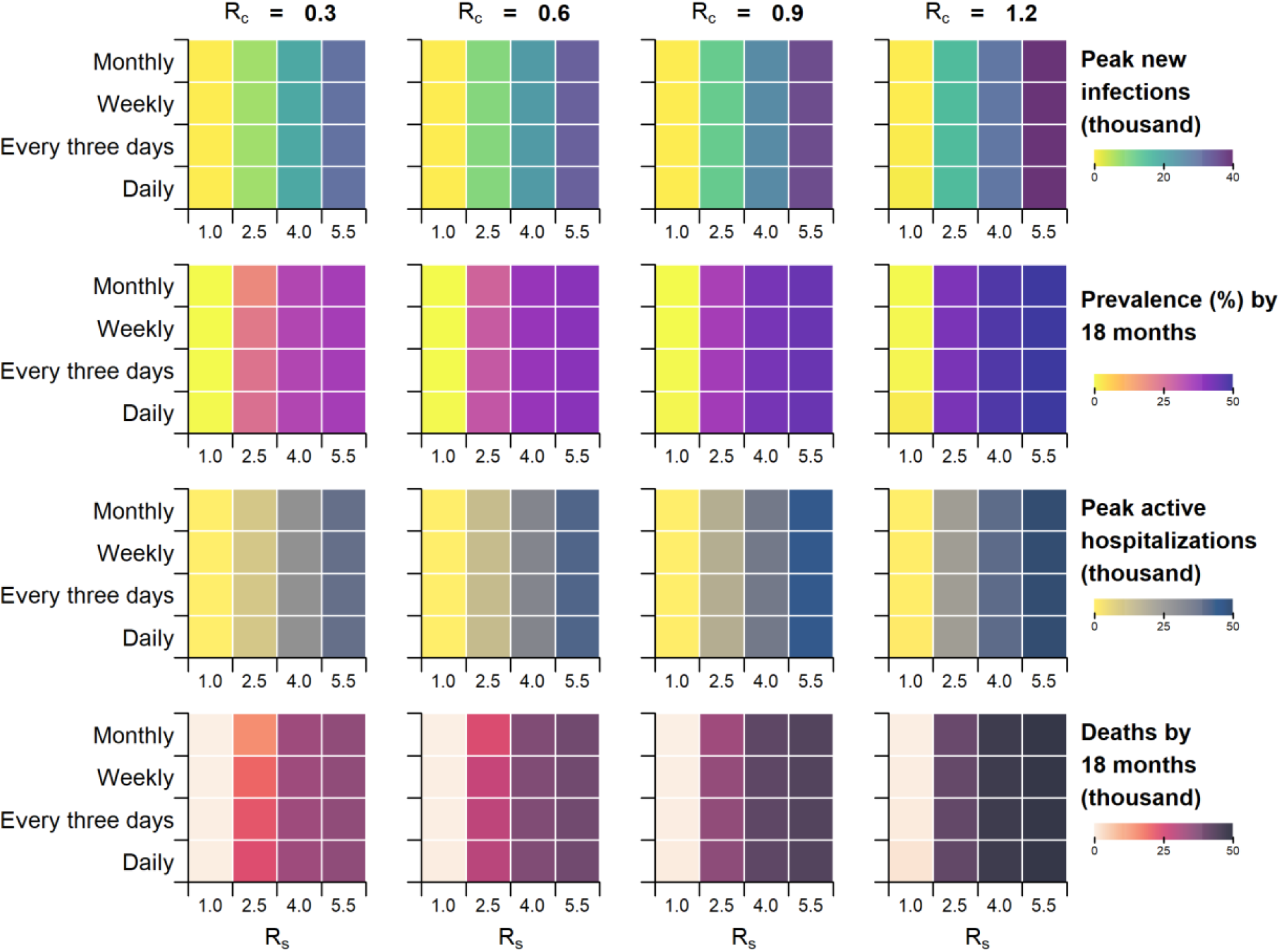
Disease burden and HCRU for outbreaks with varying importation frequencies and disease transmissibility. The indicators include peak number of new local infections (Row 1), prevalence (i.e., proportion of infections) in the general local population by the end of 18 months following the initial importation event (Row 2), peak number of in-patient mpox infections in normal wards (Row 3), and total number of deaths within 18 months following the initial importation event (Row 4).

### 3.3 Influence of NPIs on outbreak control

The simulations demonstrated the substantial positive effects of NPIs on managing the hypothetical mpox outbreak. Isolation of diagnosed cases, which was expected to halve the infectivity, would reduce the outbreak scale by 34.7%, with the peak number of new infections falling below 14,000. The timing of the outbreak peak was also postponed by two months compared to the scenario without interventions. Adding pre-departure screening for international arrivals would produce only a marginal decrease of 0.04 percentage points in infection size, but the time for the peak of new infections was further delayed by one month. The outbreak timeline was extended further with the implementation of contact tracing alongside the aforementioned two NPIs. In this scenario, a surge in infections was projected to occur 10 months after the initial importation, with the epidemic peak reached by 15 months, while the peak new infection size was additionally lowered by 1.1 percentage points. The influence on HCRU mirrored the effect on outbreak size for each NPI strategy (Figure 4, Table S8).

**Figure 4.**
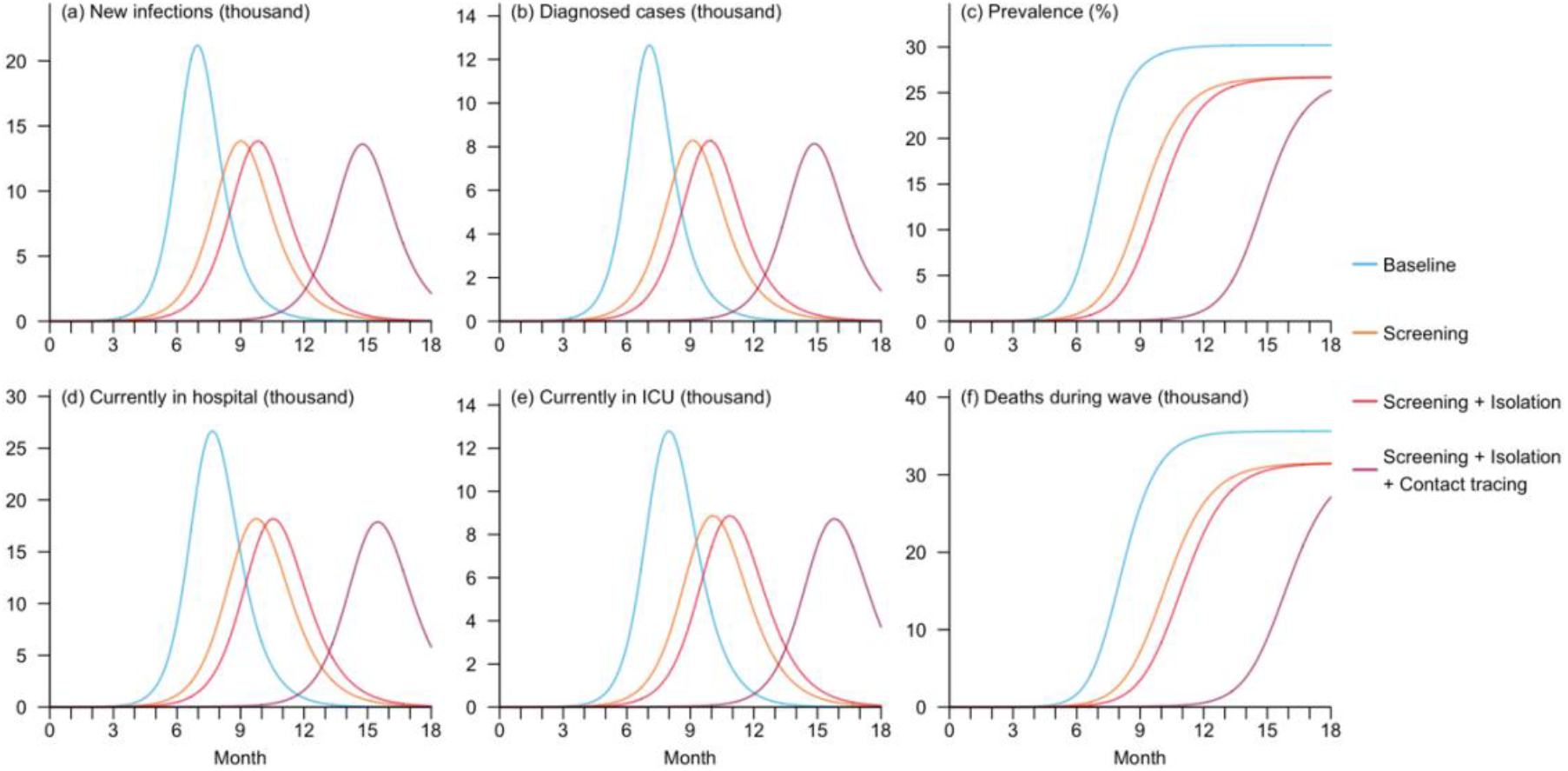
Disease burden and HCRU during mpox outbreaks for hypothetical mpox outbreaks in the scenarios with no intervention or at least one NPI implemented. The indicators include number of new local infections (a) and diagnosed cases (b) on each day, disease prevalence in the overall local population (c), number of hospitalized cases in normal wards (d) and ICU occupancy (e), as well as cumulative number of deaths (f) over the 18 months following the initial importation event.

### 3.4 Effects of vaccination on morbidity and mortality reduction

In addition to the NPIs, vaccination also effectively reduced disease morbidity and mortality. Compared to the baseline scenario, an increase in vaccination coverage of 5% in the overall population would reduce the peak infection size by 30.1%, 14.9%, and 23.2%, while preventing 16.4%, 7.7%, and 12.7% deaths, respectively, under the three vaccine allocation strategies we proposed (Figure S2). Furthermore, prioritising vaccination among the sexually active subpopulation would result in the greatest efficacy in reducing infection and fatality size across all scenarios. The distinction was the more pronounced in settings where *R*_*s*_ was neither too high nor too low across scenarios with different *R*_*c*_’s. For instance, in the setting with *R*_*c*_ = 0.6, the proportion of infections averted by the best allocation strategy, V1 (i.e., all the vaccines administered to the sexually active group), was 1.4 times that of the second optimal strategy, V3 (i.e., higher vaccine uptake in the sexually active group) when *R*_*s*_ = 2.5, while the corresponding ratio was 1.3 when *R*_*s*_ = 5.5. Furthermore, the effectiveness of vaccination on preventing infections or deaths did not consistently increase with higher disease transmissibility of either sexual or non-sexual transmission, as demonstrated by in scenarios with *R*_*s*_ ≥ 2.5 or *R*_*c*_ ≥ 0.9 in our simulations (Figure 5).

**Figure 5.**
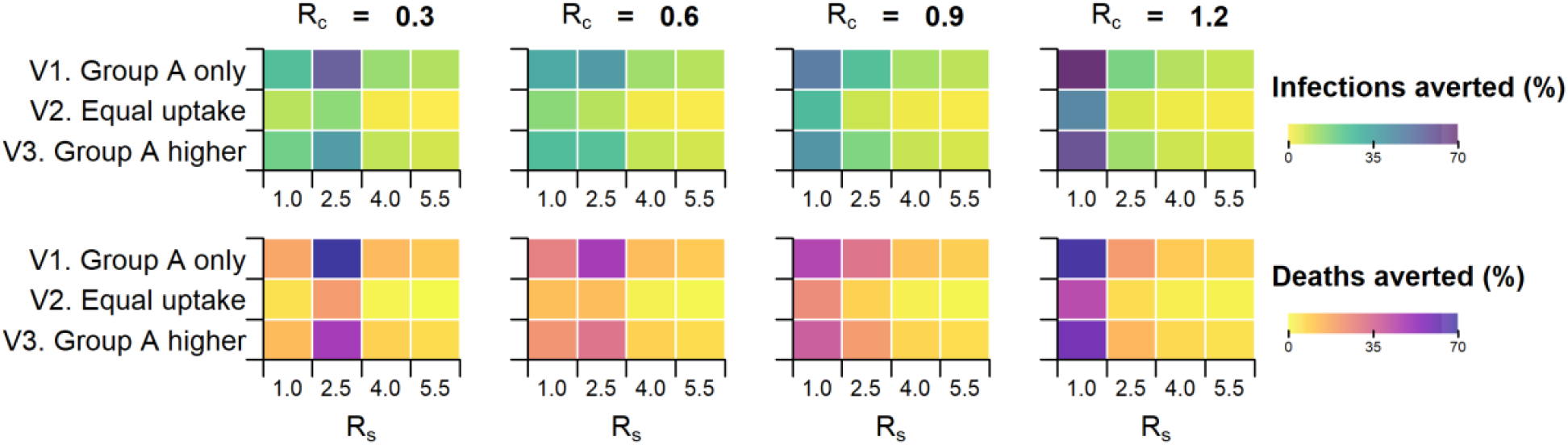
Reduction in disease burden by the three proposed vaccination strategies. Group A refers to the sexually active subpopulation. The measures to quantify the decrease include the proportion of local infections (Row 1) and deaths (Row 2) averted compared to scenarios without vaccination over the 18 months following the initial importation event.

## 4. Discussion

In this study, we simulated mpox outbreaks under various hypothetical scenarios to predict the potential spread of Clade Ib MPXV in Singapore, and assessed the impacts of vaccination and NPIs inclusive of case isolation, quarantining of close contacts and contact tracing. Our simulations revealed a high possibility of large-scale transmission with steep epidemic curves, especially in scenarios where the virus was highly transmissible through sexual routes. In the scenario with moderate disease transmissibility through non-sexual means and when the general population, rather than specific subpopulations, were affected, outbreaks would result in 30% and 25% of the total population being infected with and without control through NPIs, respectively.

Nevertheless, at least a one-month delay was observed from the initial importation event before exponential growth in infections across various scenarios assessed. This could provide decisionmakers with sufficient time to set up emergency infrastructure, testing centres and create treatment guidelines, and to also then begin containing any potential spread before the number of infections become a burden on the healthcare system. In addition, mandatory home isolation of all diagnosed cases, covering 60% of local infections, was projected to reduce the peak number of new infections by 35% and the cumulative infection size by 11%. While incomplete adherence could lead to secondary infections of close contacts, particularly if non-sexual transmission be an established pathway of transmission [6], isolation at home remains more feasible than in healthcare settings. Given the much longer infectious period of mpox compared to COVID-19 [18]. extended isolation could easily strain pre-existing infrastructure with the surge in case counts.

Although pre-departure testing was shown to be effective in preventing importation during the COVID-19 pandemic [19]. its limited effectiveness in our simulations might be explained by the high transmissibility of the MPXV through sexual contact, which resulted in rapid local disease spread, thereby overshadowing the role of importation in driving the outbreak. In contrast, the extra benefits of quarantining close contacts beyond isolation and screening was projected to be much greater in delaying the outbreak, reflecting its extensive use during the COVID-19 pandemic [20,21]. Nevertheless, it is important to note that contact tracing would require substantial human resources for identifying the close contacts, which may not always be available. Isolation is therefore recommended as the primary intervention strategy with the addition of screening or contact tracing as additional measures. In the ongoing mpox outbreak of 2024, most identified patients infected with Clade Ib MPXV outside Africa were imported cases with travel history from high-risk countries [22], making it plausible that any mpox case outbreaks in APAC would be driven by importation with minimal local spread. However, as demonstrated by our simulations, it is also possible that an outbreak could occur depending on mpox transmissibility (Figure 3). We therefore examined the proposed NPIs’ effects under these two scenarios in a subsequent sensitivity analysis, in which the virus’s transmissibility for non-sexual transmission were set to be nearly zero or significantly higher than the baseline scenario. The marginal differences observed in the proportion of infections and hospital admissions averted by these measures consistently demonstrated their effectiveness in containing the outbreak (Figure S7–8, Table S10–11).

While our model outputs supports the pivotal role vaccination played in controlling the epidemic, the results also emphasised the importance of its strategic implementation [23]. Prioritising the most vulnerable group, the sexually active individuals in this context, would maximise its reduction in the overall disease burden. Nonetheless, vaccination might not be optimal in our setting of an outbreak in an APAC city, as its projected benefits were no higher than those brought by various NPIs. Although these limited discrepancies in effects between the two types of interventions might be explained by the relatively low vaccination scale (5%) in our proposed strategies, NPIs could be more sustainable considering the high prices of approved mpox vaccines and the time required for them to take effect in individuals [24]. Moreover, mass vaccination campaigns often face barriers such as the limited vaccine supply and shortage of healthcare workers [25]. Another challenge may be potential vaccine hesitancy in the APAC region, driven by historic public health events, safety concerns, and spread of misinformation [26–28]. Additionally, as the 2022 mpox outbreak primarily affected the GBMSM community, the general public might perceive a low risk of infection, resulting in diminished recognition of the vaccine’s importance and hence reduced motivation in immunisation [29]. These obstacles would collectively make it unlikely to increase vaccine uptake rapidly enough to prevent a forthcoming outbreak. Vaccination for the general APAC population at this time may also result in a lack of vaccines being available for the high-risk groups, especially among countries with ongoing outbreaks, facilitating the continued mitigation of global transmission risks.

One major limitation of our study pertained to the paucity of informative statistics. Our simulated trajectories suggested that the current outbreak potentially might be in an early stage when compared to the surveillance data available at the time of this study, as importation from African countries accounted for most of the globally detected cases. This posed great challenges to extrapolating the disease transmissibility, the key parameters in our model, in an APAC setting where the contact patterns might differ significantly from Africa. To address the large uncertainties in model parameters, we performed a sensitivity analysis in which we simulated a range of outbreaks using various combinations of the basic reproduction numbers for transmission through sexual (*R*_*s*_) and non-sexual (*R*_*c*_) activities. This aimed to capture both the potential extremes caused by super-spreaders and the most probable epidemic trajectories (Figure S1). Furthermore, the predominance of the available surveillance data sourcing from Africa, where healthcare and monitoring systems might face constraints, could lead to an underestimation of infection sizes and an overestimation of disease severity. Consequently, the simulated HCRU projected from the African data in our study might far exceed reality in APAC, offering a potential silver lining. With the availability of more case data and epidemiological parameters, these models require updating.

Another limitation was the simplification of real-world scenarios in our model, such as hospital discharge and readmission of patients who had not fully recovered, as well as variations in population size due to natural birth and deaths. However, this was believed to have a minimal impact owing to the large population affected by the outbreak. We also streamlined the clinical testing and quarantine processes, assuming accurate testing results and no tests or quarantine for uninfected individuals. In addition, we did not explicitly account for international arrivals who were exposed to the disease but not yet infectious, as the lack of relevant travel data made it challenging to quantify the exact population. Nevertheless, we have built in such possibilities by adjusting sensitivity of the pre-departure screening to the relatively low 60%.

Despite these limitations, the transmission model proposed in this study is sufficiently flexible to accommodate for a range of scenarios with diverse disease transmission dynamics, enabling decisionmakers to plan for different potential outbreak possibilities. This study explores the impact of NPI impacts on mpox outbreaks, an area much less explored compared to the virus’s transmission patterns [15,30]. This has provided evidence for adapting the containment measures widely applied during the COVID-19 pandemic to effectively managing new outbreaks with probably high community transmission potential.

Our simulations suggest that while mpox outbreaks triggered by importation could lead to substantial morbidity and mortality in an APAC city with low immunity to the virus, they would still remain controllable provided adequate preparation and a timely response from decisionmakers, underscoring the significance of robust surveillance systems. In addition, the NPIs explored are able to delay and suppress outbreak sizes and curb disease spread up to 36%. Based on this, mass vaccination of the general population is not recommended but surveillance systems, infrastructure for contact tracing, quarantining of close contacts and institutional isolation of cases is urgently required.

## Data Availability

Data available upon request

## Author contributions

SJ, GG, and BLD conceived and designed the study. SJ and GG implemented the statistical analysis and created the figures and tables. SJ wrote the original draft of the manuscript. GG, AK, KP, RKJT, JTL, and BLD reviewed and edited the manuscript.

