## Supplementary for "Modelling the potential spread of Clade Ib MPXV in an Asia Pacific city"

**Supplementary Information**

### Additional Explanations for the Model and Proposed Scenarios

**Table S1. Name, definition, and derivation approach (where appliable) of the model parameters.**

| **Parameter** | **Definition** | **Derivation approach** |
| --- | --- | --- |
| *Population-related* |  |  |
| $N$ | Overall population size |  |
| $p_{A}$ | Proportion of sexually active individuals in the general population |  |
| $p_{L}$ | Proportion of sexually inactive individuals in the general population |  |
| $N_{A}$ | Sexually active population size | $p_{A}N$ |
| $N_{L}$ | Sexually inactive population size | $p_{L}N$ |
| *Transmission-related* |  |  |
| $R_{s}$ | Average number of secondary infections generated by an infector through sexual routes in a population with no previous exposure to the disease |  |
| $R_{c}$ | Average number of secondary infections generated by an infector through non-sexual routes in a population with no previous exposure to the disease (i.e., in the general community) |  |
| $\beta_{s}$ | Rate of disease transmission through sexual activities | $\gamma_{I}R_{s}/N_{A}$ |
| $\beta_{c}$ | Rate of disease transmission through non-sexual activities | $\gamma_{I}R_{c}/(N_{A}+N_{L})$ |
| $c_{iso}$ | Proportion of reduction in the number of secondary infections due to quarantine or isolation |  |
| $\alpha$ | Rate of latent individuals becoming infectious | Inverse of average duration from initial exposure to onset of infectiousness |
| $\gamma_{I0}$ | Recovery rate for the undiagnosed infectious | Inverse of average recovery period |
| $\gamma_{II}$ | Recovery rate for the diagnosed cases not requiring hospitalization | Inverse of the difference between average recovery period and duration from onset of infectiousness to diagnosis |
| $\gamma_{HI}$ | Hospitalization rate among infectious cases to be hospitalized | Inverse of the difference between average duration from infectious to hospitalization and duration from onset of infectiousness to diagnosis |
| $\gamma_{HH}$ | Recovery rate among hospitalized infectious cases not to be transferred to Intensive Care Unit (ICU) | Inverse of the average duration from hospitalization to recovery (for patients not transferred to ICU) |
| $\gamma_{UH}$ | ICU admission rate among hospitalized cases to be transferred to ICU | Inverse of the average duration from hospitalization to ICU admission |
| $\gamma_{UU}$ | Recovery from ICU | Inverse of the average duration from ICU admission to recovery |
| $\gamma_{DU}$ | Death rate | Inverse of the average duration from  ICU admission to death |
| *Severity-related* |  |  |
| $p_{H}$ | Case-hospitalization risk | Ratio between confirmed and hospitalized cases, excluding those to be admitted to ICU |
| $p_{I}$ | Case-ICU risk | Ratio between confirmed cases and ICU admissions, excluding those who would eventually die |
| $p_{D}$ | Case-fatality risk | Ratio between confirmed cases and deaths |
| *Surveillance-related* |  |  |
| $dr$ | Diagnostic rate | Proportion of infections who were clinically diagnosed (and thus reported as cases) |
| $dr_{imp}$ | Diagnostic rate for importation | Proportion of imported infections who were clinically diagnosed (and thus reported as cases) |
| $\phi_{I}$ | Rate of notification | Inverse of average duration from clinical testing to diagnosis |
| $ct_{max}$ | Maximum number of new cases to perform contact tracing per day |  |
| $ct_{connect}$ | Average number of close contacts traced per new case |  |
| $N_{CT}$ | Number of traced close contacts | Capped at the minimum of exposed population and $(ct_{max}\cdot ct_{connect})$ |
| $\sigma$ | Pre-departure testing sensitivity | Proportion of infections who were correctly identified by pre-departure tests |
| *Vaccine-related* |  |  |
| $ve$ | Vaccine efficacy |  |
| $vacov_{A}$ | Vaccine coverage in the sexually active population |  |
| $vacov_{L}$ | Vaccine coverage in the sexually inactive population |  |

**Transition rates between different compartments of the Model**

Compartments in our model encompass Susceptible ($S_{A}$ for sexually active group and $S_{L}$ for sexually inactive group), Exposed ($E$), Quarantined ($Q)$, Infectious ($I_{u}$ for the undiagnosed, $I_{d}$ for those who had been tested but yet to be confirmed, and $C$for the diagnosed), Hospitalised ($H$), Admitted to ICU ($U$), Recovered ($R_{I}$ for recovery from home, $R_{H}$ for recovery after hospitalisation, and $R_{U}$ for recovery after ICU admission), and Dead ($D$). In addition, compartments making up of the infectious population, including $I_{u}$, $I_{d}$, and $C$, were further divided into the sexually active and inactive groups, which were denoted by subscripts $A$ and $L$ respectively. From these groups, we derived the number of infectious sexually active or inactive individuals, as $I_{A}=I_{uA}+(1-c_{iso})(I_{dA}+C_{A})$ and $I_{L}=I_{uL}+(1-c_{iso})\left( I_{dL}+C_{L} \right)$, respectively. The transition between different compartments for local transmission can be expressed as below:

$$\frac{dS_{A}}{dt}=-\left( \beta_{S}I_{A}+\beta_{C}\left( I_{A}+I_{L} \right) \right) S_{A}$$

$$\frac{dS_{L}}{dt}= -\beta_{C}\left( I_{A}+I_{L} \right)S_{L}$$

$$\frac{dE}{dt}=\left( \beta_{S}I_{A}+\beta_{C}\left( I_{A}+I_{L} \right) \right) S_{A} +\beta_{C}\left( I_{A}+I_{L} \right)S_{L} -N_{CT}-\alpha E$$

$$\frac{dQ}{dt}=N_{CT}-\alpha Q$$

$$\frac{dI_{u}}{dt}=\alpha E\cdot\left( 1-dr \right)- \gamma_{I0}I_{u}$$

$$\frac{dI_{d}}{dt}=\alpha E\cdot dr+\alpha Q- \phi_{I}I_{d}$$

$$\frac{dC}{dt}= \phi_{I}I_{d}-\gamma_{II}\left( 1-p_{H} \right)C-\gamma_{HI}p_{H}C$$

$$\frac{dH}{dt}= \gamma_{HI}p_{H}C-\gamma_{HH}\frac{p_{H}-p_{U}}{p_{H}}H-\gamma_{UH} \frac{p_{U}}{p_{H}}H$$

$$\frac{dU}{dt}= \gamma_{UH} \frac{p_{U}}{p_{H}}H- \gamma_{UH} \frac{p_{U}-p_{D}}{p_{U}}U- \gamma_{UU} \frac{p_{D}}{p_{U}}U$$

$$\frac{dR_{I}}{dt}= \gamma_{I0}I_{u}+ \gamma_{II}\left( 1-p_{H} \right)C$$

$$\frac{dR_{H}}{dt}= \gamma_{HH}H$$

$$\frac{dR_{U}}{dt}= \gamma_{UH} \frac{p_{U}-p_{D}}{p_{U}}U$$

$$\frac{dD}{dt}= \gamma_{UU} \frac{p_{D}}{p_{U}}U$$

Furthermore, infectious importation ($I^{imp}$), when present, would be categorized into subgroups $I_{uA}^{imp}$, $I_{dA}^{imp}$, $I_{uL}^{imp}$, and $I_{dL}^{imp}$ according to the diagnostic rate, $dr_{imp}$, and proportion of sexually active individuals in the affected population, i.e., $p_{A}/(p_{A}+p_{L})$. These subgroups would progress (e.g., getting tested or recovering without testing) in the same way as the local population.

**Table S2. Fixed model parameters and the rationales behind their selection.**

| **Parameter** | **Value** | **Rationale/Reference** |
| --- | --- | --- |
| Overall population size, $N$ | 5,800,000 | Statistics derived from numbers published by the Singapore government [1] |
| Average incubation period | 7d | Statistics derived from literature [2] |
| Average duration from clinical testing to diagnosis | 3d | Statistics derived from literature [3] |
| Average duration from being infectious to hospitalization | 7d | Assumption made in the absence of relevant data: Infectious individuals experiencing worsening symptoms or health complications were assumed to seek in-patient care approximately one week after becoming infectious |
| Average duration from hospitalization to ICU admission | 7d | Assumption made in the absence of relevant data: Hospitalized patients with mpox who failed to recover in a regular ward would be transferred to ICU |
| Average recovery period for home-recovery cases | 21d | Statistics derived from literature [4] |
| Average duration from hospitalization to recovery (for patients not admitted to ICU) | 7d | Statistics derived from literature [5] |
| Average duration from ICU admission to recovery | 10d | Assumption made in the absence of relevant data |
| Average duration from ICU admission to death | 10d | Assumption made in the absence of relevant data |
| Case-hospitalization risk, $p_{H}$ | 10% | Statistics derived from literature [6] |
| Case-ICU risk, $p_{U}$ | 3.5% | Statistics derived from literature [6] |
| Case-fatality risk, $p_{D}$ | 1.0% | Statistics derived from literature [6] |
| Diagnostic rate, $dr$ | 60% | Undiagnosed infections include those asymptomatic and those who not seeking medical assistance |
| Diagnostic rate for importation, $dr_{imp}$ | 30% | Undiagnosed infections include those asymptomatic and those who not seeking medical assistance |

**Table S3. Adjustable parameter values across the five baseline scenarios.**

| **Parameter** | **Scenario 1** | **Scenario 2** | **Scenario 3a** | **Scenario 3b** | **Scenario 3c** |
| --- | --- | --- | --- | --- | --- |
| Proportion of sexually active individuals in the general population, $p_{A}$^#^ | 4.6% | 5.7% | 26% | 26% | 26% |
| Proportion of sexually inactive individuals in the general population, $p_{L}$^#^ | 1.9% | 0.6% | 39% | 39% | 39% |
| Average number of secondary infections generated by an infector through sexual routes in a population with no previous exposure to the disease, $R_{S}$^#^ | 9.4 | 6.4 | 4.0 | 4.0 | 4.0 |
| Average number of secondary infections generated by an infector through non-sexual routes in a population with no previous exposure to the disease, $R_{C}$^*^ | 0.6 | 0.6 | 0.3 | 0.6 | 0.9 |

^#^ Assumption made based on unpublished data in Singapore.
^*^ Assumption made in the absence of relevant data.

**Explanations for individual non-pharmaceutical interventions (NPIs)**

*Isolation*

All infections who underwent clinical testing were assumed to start quarantine immediately after testing and transit to home isolation upon being confirmed as a mpox case. Nevertheless, diagnosis and reporting were delayed by approximately three days due to the time required for laboratory tests. Since quarantine and isolation occurred at home, there was a risk of household transmission and non-compliance. Consequently, we did not completely eliminate the risk of transmission and assumed a 50% reduction in the average number of secondary infections generated by quarantined or isolated infections. In scenarios without this intervention, the relevant parameter, $c_{iso}$, was set to 50% to account for the reduced contact between the susceptible population and the infectious who sought medical attention, allowing for the possibility of self-isolation.

*Contact tracing*

The number of traced close contacts ($N_{CT}$) on day $t$ was determined by the number of diagnosed cases on day $(t-1)$ and exposed but yet infectious individuals before day $t$, contact tracing capacity, as well as the number of close contacts per case (Table S1). We did not consider mandatory testing for individuals in quarantine, thereby applying the same diagnostic rate and risks of developing severe symptoms (i.e., getting hospitalized, admitted to ICU, or dying) to both the traced close contacts and the unquarantined local infections. In scenarios without this intervention, $N_{CT}=ct_{max}=0$.

*Screening*

International arrivals who had visited high-risk territories within 21 days prior to departure would only be permitted entry the border with negative test results obtained within three days pre-departure. Activity restrictions were also required between testing and departure to ensure relatively high sensitivity of pre-departure test. The sensitivity parameter, $\sigma$, was assumed to be 60% to account for undetected infections or individuals exposed to the virus but not yet infectious (Table S4). This pre-departure screening would lead to a $100(1-\sigma)\%$ reduction in the number of imported infections per importation event. In scenarios where *isolation* is implemented for local infections, the imported infections who sought medical assistance post-arrival were also expected to quarantine or isolate themselves in designated facilities, such as hotels, upon testing.

**Table S4. Parameter values for individual NPIs.**

| **NPI** | **Parameters involved** | **Values when intervention is implemented** |
| --- | --- | --- |
| *Community-level* |  |  |
| Isolation | Proportion of reduction in the number of secondary infections due to quarantine or isolation, $c_{iso}$ | 75% |
| Contact tracing | Maximum number of new cases to perform contact tracing per day, $ct_{max}$  Average number of close contacts traced per new case, $ct_{connect}$ | 7  10 |
| *Border-level* |  |  |
| Screening | Pre-departure testing sensitivity, $\sigma$ | 60% |

**Table S5. Hypothetical vaccination uptake rates for the sexually active and inactive groups.**

| **Allocation plan** | **Sexually active group** | **Sexually inactive group** |
| --- | --- | --- |
| V1. Sexually active group only | 19.2% | 0.0% |
| V2. Equal uptake | 7.7% | 7.7% |
| V3. Sexually active group higher | 14.4% | 3.2% |

### Simulation Results

**Table S6. Ratio of four disease burden and healthcare resource utilization (HCRU) indicators between the baseline scenario (denominator) and scenarios with varying disease transmissibility but a fixed importation frequency of five per three days (numerator).**

| $\boldsymbol{R}_{\boldsymbol{C}}$ | $\boldsymbol{R}_{\boldsymbol{S}}$ | **Peak new infections** | **Prevalence by 18 months** | **Peak active hospitalizations** | **Deaths by 18 months** |
| --- | --- | --- | --- | --- | --- |
| 0.3 | 1.0 | 0.0001 | 0.0007 | 0.0002 | 0.001 |
|  | 2.5 | 0.28 | 0.62 | 0.33 | 0.59 |
|  | 4.0 | 0.72 | 0.88 | 0.88 | 0.88 |
|  | 5.5 | 1.15 | 0.94 | 1.25 | 0.94 |
| 0.6 | 1.0 | 0.0004 | 0.002 | 0.0004 | 0.002 |
|  | 2.5 | 0.41 | 0.78 | 0.44 | 0.76 |
|  | 4.0 | 1.00 | 1.00 | 1.00 | 1.00 |
|  | 5.5 | 1.48 | 1.02 | 1.45 | 1.06 |
| 0.9 | 1.0 | 0.001 | 0.005 | 0.002 | 0.004 |
|  | 2.5 | 0.54 | 0.94 | 0.58 | 0.93 |
|  | 4.0 | 1.14 | 1.14 | 1.14 | 1.14 |
|  | 5.5 | 1.62 | 1.19 | 1.50 | 1.19 |
| 1.2 | 1.0 | 0.01 | 0.02 | 0.01 | 0.02 |
|  | 2.5 | 0.70 | 1.12 | 0.74 | 1.12 |
|  | 4.0 | 1.30 | 1.28 | 1.29 | 1.28 |
|  | 5.5 | 1.77 | 1.33 | 1.64 | 1.33 |

**Table S7. Ratio of four disease burden and HCRU indicators between the scenario with an importation frequency once per three days (denominator) and scenarios with other importation frequencies (numerator), when** $\boldsymbol{R}_{\boldsymbol{S}}\boldsymbol{=1}\boldsymbol{.}\boldsymbol{0}$ **and** $\boldsymbol{R}_{\boldsymbol{C}}\boldsymbol{=0}\boldsymbol{.}\boldsymbol{9}$**.**

| **Importation Frequency** | **Peak Infections** | **Prevalence by 18 months** | **Peak active Hospitalisation** | **Deaths by**  **18 months** |
| --- | --- | --- | --- | --- |
| Monthly | 0.10 | 0.10 | 0.10 | 0.11 |
| Weekly | 0.43 | 0.43 | 0.44 | 0.43 |
| Daily | 2.89 | 2.94 | 2.88 | 2.96 |

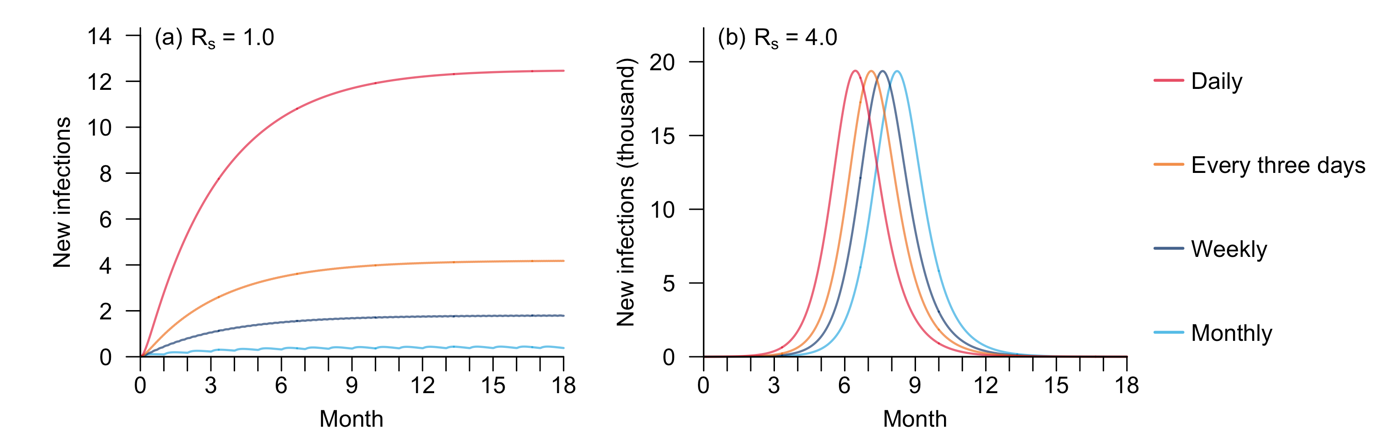

**Figure S1. Number of new local infections over the 18 months following the initial importation event in scenarios with diverse importation frequencies, when** $\boldsymbol{R}_{\boldsymbol{c}}\boldsymbol{=0}\boldsymbol{.}\boldsymbol{4}$ **and** $\boldsymbol{R}_{\boldsymbol{s}}\boldsymbol{=1}\boldsymbol{.}\boldsymbol{0}$ **(a) or** $\boldsymbol{4}\boldsymbol{.}\boldsymbol{0}$ **(b).**

**Table S8. Disease burden and HCRU indicators for hypothetical mpox outbreaks in the baseline scenario and scenarios with NPIs implemented.** Peak number of new infections and time with maximum number of new infections were calculated for local infections only, while peak number of active hospitalizations (normal ward) and time with maximum hospital occupancy were reported for the overall population.

| **Scenario** | **Peak Infections**  **(n)** | **Time when new infections peaked (month)** | **Peak active hospitalisations**  **(n)** | **Time when hospital occupancy peaked (month)** |
| --- | --- | --- | --- | --- |
| Baseline | 21,204 | 7.0 | 26,643 | 7.7 |
| Isolation | 13,844 | 9.0 | 18,180 | 9.7 |
| Isolation +  Screening | 13,836 | 9.8 | 18,170 | 10.5 |
| Isolation +  Screening +  Contact tracing | 13,599 | 14.8 | 17,877 | 15.5 |

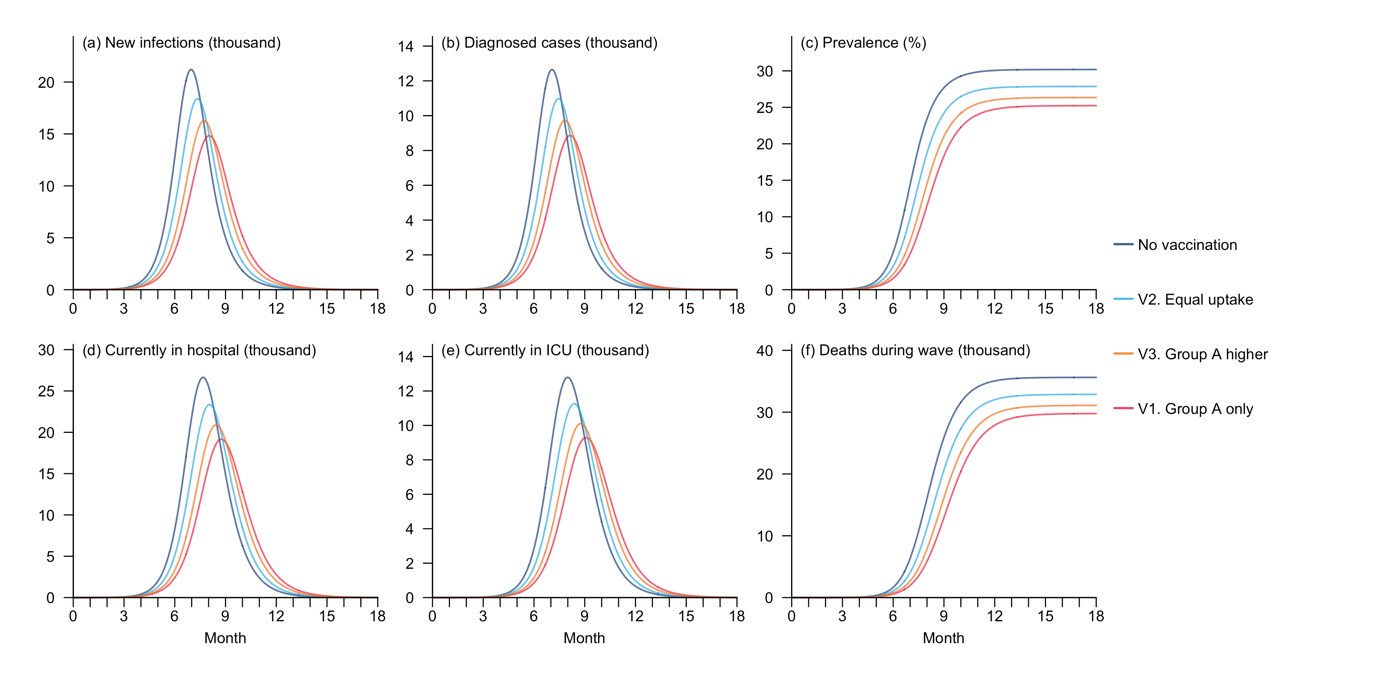

**Figure S2. Disease burden and HCRU during mpox outbreaks for hypothetical mpox outbreaks in the baseline scenario and scenarios with vaccination (where** $\boldsymbol{R}_{\boldsymbol{s}}\boldsymbol{=4}\boldsymbol{.}\boldsymbol{0}$ **and** $\boldsymbol{R}_{\boldsymbol{c}}\boldsymbol{=0}\boldsymbol{.}\boldsymbol{6}$**).** Group A refers to the sexually active subpopulation. The indicators include number of new local infections (a) and diagnosed cases (b) on each day, disease prevalence in the overall local population (c), number of hospitalized cases in normal wards (d) and ICU occupancy (e), as well as cumulative number of deaths (f) over the 18 months following the initial importation event.

### Sensitivity Analysis: Epidemic trajectories in scenarios with different disease transmissibility

Given the great uncertainty in the transmissibility of Clade Ib MPXV at the time of this study, we conducted a sensitivity analysis to explore potential scales of mpox outbreak should the $R_{0}$ for sexual or non-sexual transmission diverge from the setting in our baseline scenario.

Specifically, we modelled $R_{s}$ and $R_{c}$ as random variables, with $R_{s}$ following a right-skewed gamma distribution (mean = 4.00 and standard deviation (SD) = 1.14), which was developed from the results of our ongoing survey on sexual behaviours in Singapore, to account for the potential presence of superspreaders, and $R_{c}$ characterised by a normal distribution (mean = 0.60, SD = 0.24), as super-spreading was less common in transmission through non-sexual means. We then randomly sampled 501 pairs of $R_{s}$ and $R_{c}$ from their respective distributions and simulated outbreaks utilising these transmissibility values, keeping other model parameters same as those in the baseline scenario. Trajectories for all the simulated outbreaks are visualised in Figure S3, along with the median run highlighting the curves for the outbreak with the median overall infection size by 18 months.

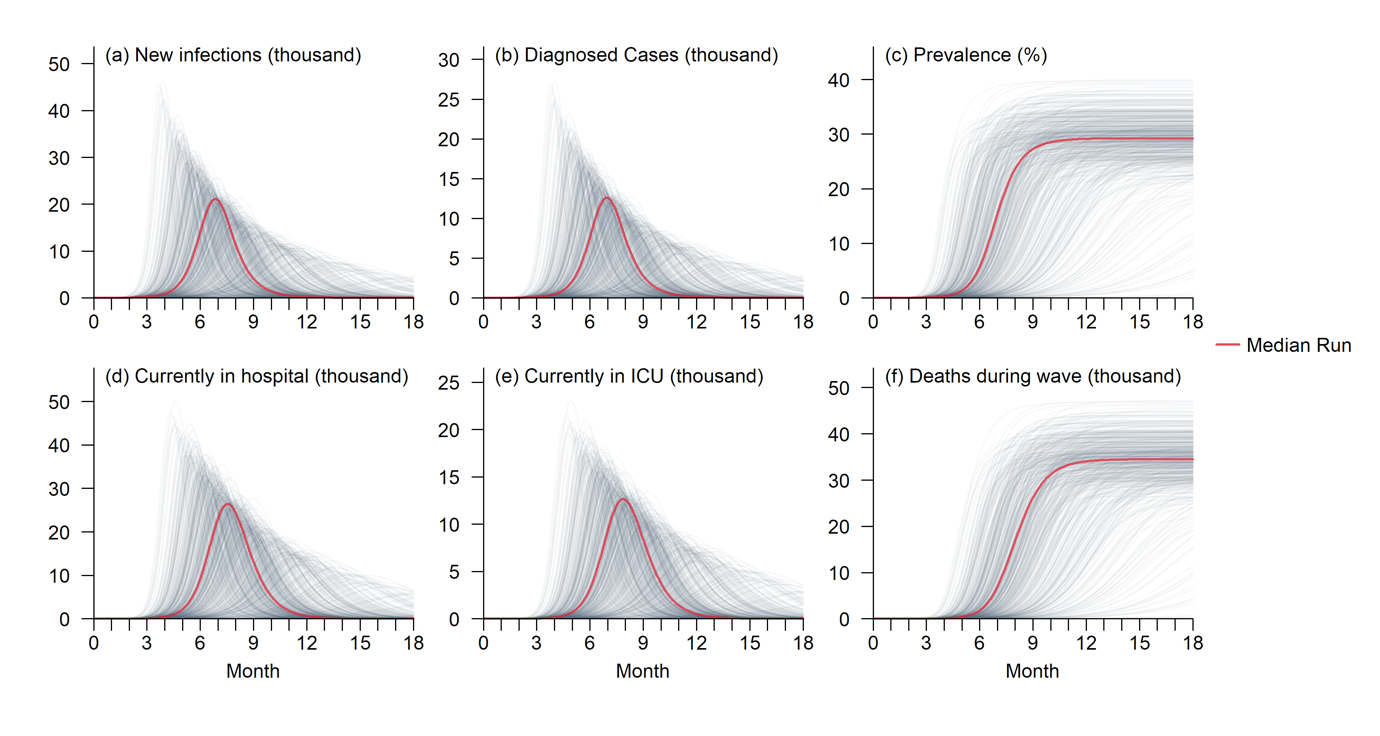

**Figure S3. Disease burden and HCRU with varying levels of disease transmissibility.** The indicators include number of new local infections (a) and diagnosed cases (b) on each day, disease prevalence in the overall local population (c), number of hospitalized cases in normal wards (d) and ICU occupancy (e), as well as cumulative number of deaths (f) over the 18 months following the initial importation event.

### Sensitivity Analysis: Disease spread restricted to specific subpopulations

A sensitivity analysis was performed to predict the epidemic trajectories when the transmission of Clade Ib monkeypox virus (MPXV) was confined to specific high-risk subpopulations and their close contacts, but with reduced transmissibility of sexual transmission. Specifically, we adapted Scenario 1 and 2 to simulate outbreaks driven by the men-who-have-sex-with-men (GBMSM) community and sex workers in Singapore, respectively, using the transmissibility parameter pair for the baseline scenario, i.e., $R_{s}=4.0$ and $R_{c}=0.6$. This would lead to a smaller and delayed wave in both scenarios (Figure S4).

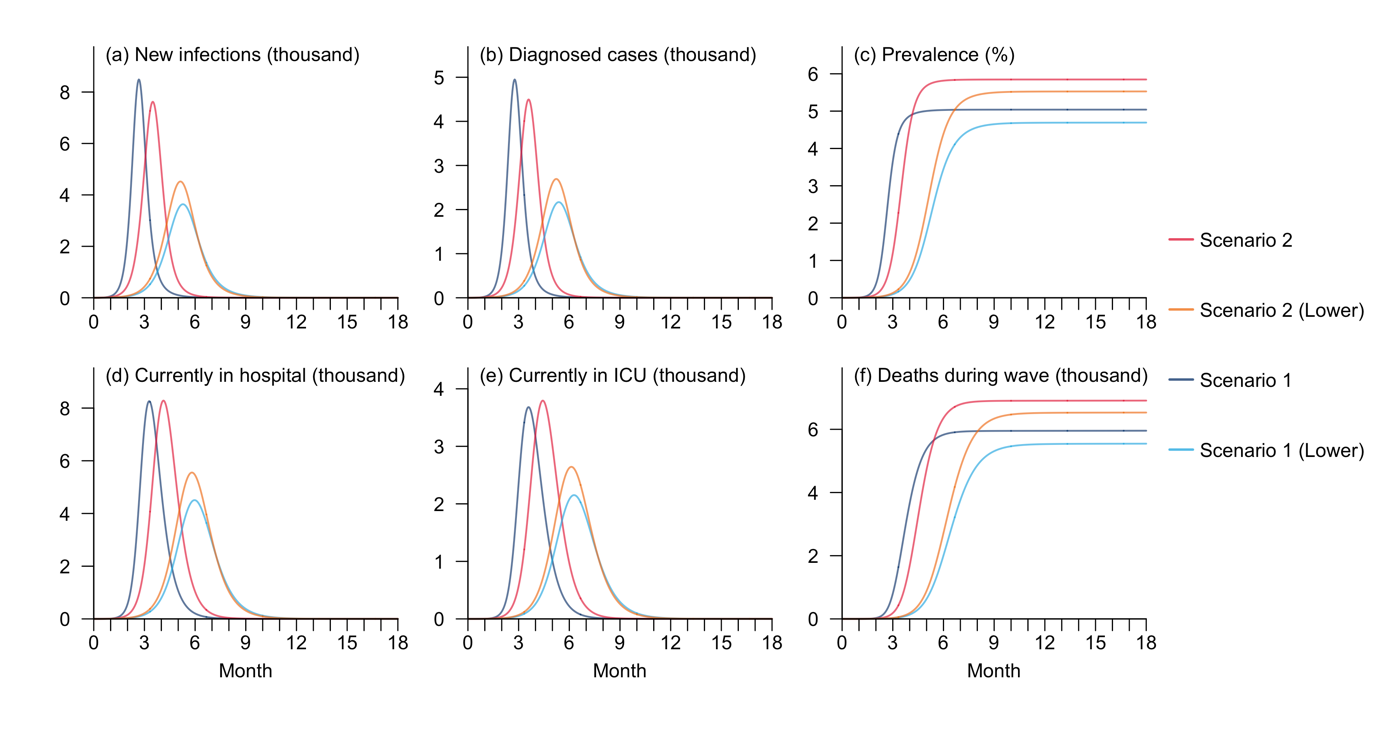

**Figure S4. Disease burden and HCRU with different levels of disease transmissibility for Scenario 1 and 2.** ‘Lower’ refers to scenarios with $R_{s}=4.0$, which is lower than the setting in Scenario 1 or 2. The indicators include number of new local infections (a) and diagnosed cases (b) on each day, disease prevalence in the overall local population (c), number of hospitalized cases in normal wards (d) and ICU occupancy (e), as well as cumulative number of deaths (f) over the 18 months following the initial importation event.

### Sensitivity Analysis: Impacts of self-isolation on transmission patterns

One assumption of our model is that infections who underwent clinical testing tended to reduce their contacts (i.e., self-isolate) with others even in the absence of mandatory home isolation. In the main analysis we presumed the reduction as 50%. Nevertheless, the degree of self-isolation might vary by individual. We therefore performed a sensitivity analysis was performed to assess the impacts of self-isolation on transmission dynamics. Specifically, we considered four scenarios, with the contact reductions of 0%, 25%, 50%, and 75%, respectively, while keeping other parameters same as those of the baseline scenario. No additional interventions were considered in this analysis. This reduction in contact would lead to substantial decrease in the outbreak size and the burden on healthcare systems, as well as a delay in the outbreak timeline (Figure S5).

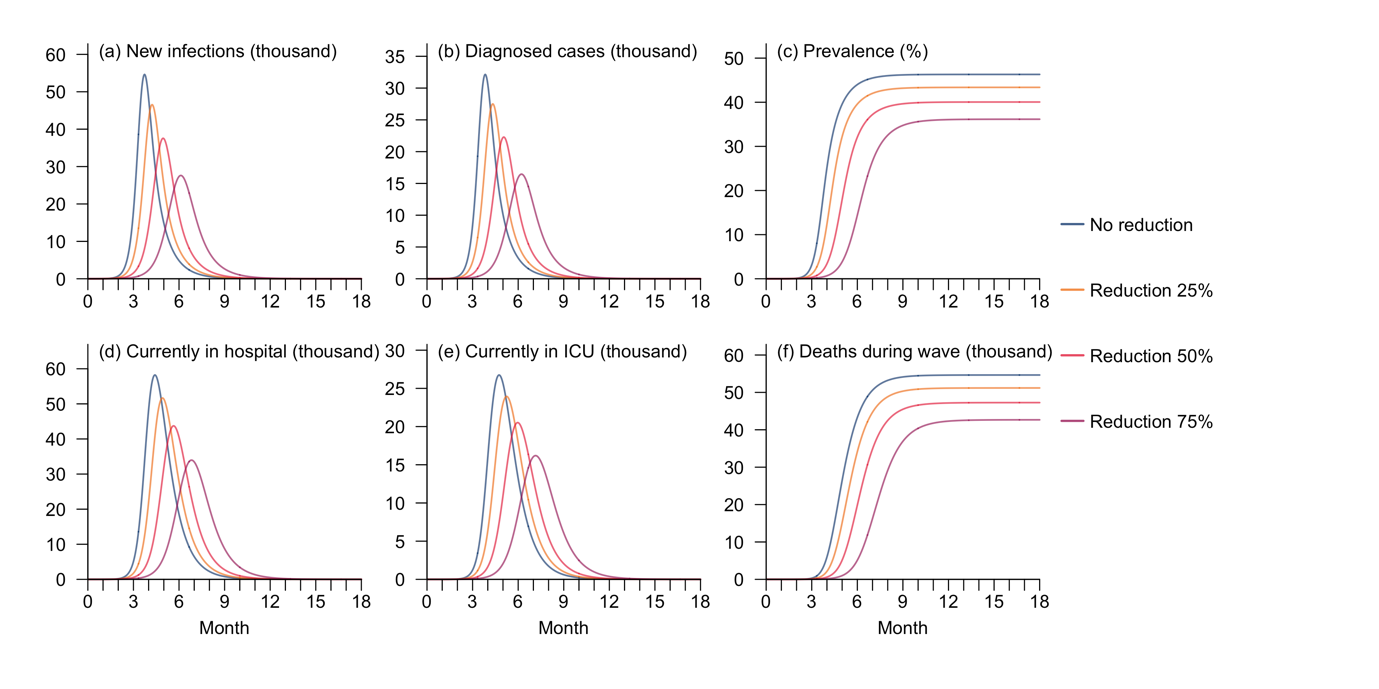

**Figure S5. Disease burden and HCRU with varying degrees of reduction in contacts due to self-isolation under the baseline scenario.** The indicators include number of new local infections (a) and diagnosed cases (b) on each day, disease prevalence in the overall local population (c), number of hospitalized cases in normal wards (d) and ICU occupancy (e), as well as cumulative number of deaths (f) over the 18 months following the initial importation event.

### Sensitivity Analysis: Impacts of pre-departure testing sensitivity on intervention effectiveness

In the main analysis we assumed a pre-departure testing sensitivity of 60%, meaning 60% of the infectious travellers from abroad would be prevented from entering this country. However, this sensitivity parameter value depends highly on factors such as reliability of testing kits and risk of infection prior to departure (which would ideally be zero if the activity restrictions were stringent). To assess the influence of testing sensitivity on effectiveness of screening, we conducted a sensitivity analysis, comparing the projected epidemic trajectories under scenarios with no testing and with pre-departure testing sensitivity ranging from 20% to 80%. For consistency, home isolation was assumed to be in place in all the scenarios included in this analysis. Although the reduction in outbreak size attributed to the raise in testing sensitivity was hardly observable, a mild delay in outbreak timeline was observed as the sensitivity of pre-departure screening increased, with the duration of postponement positively correlated with the sensitivity value (Figure S6, Table S9).

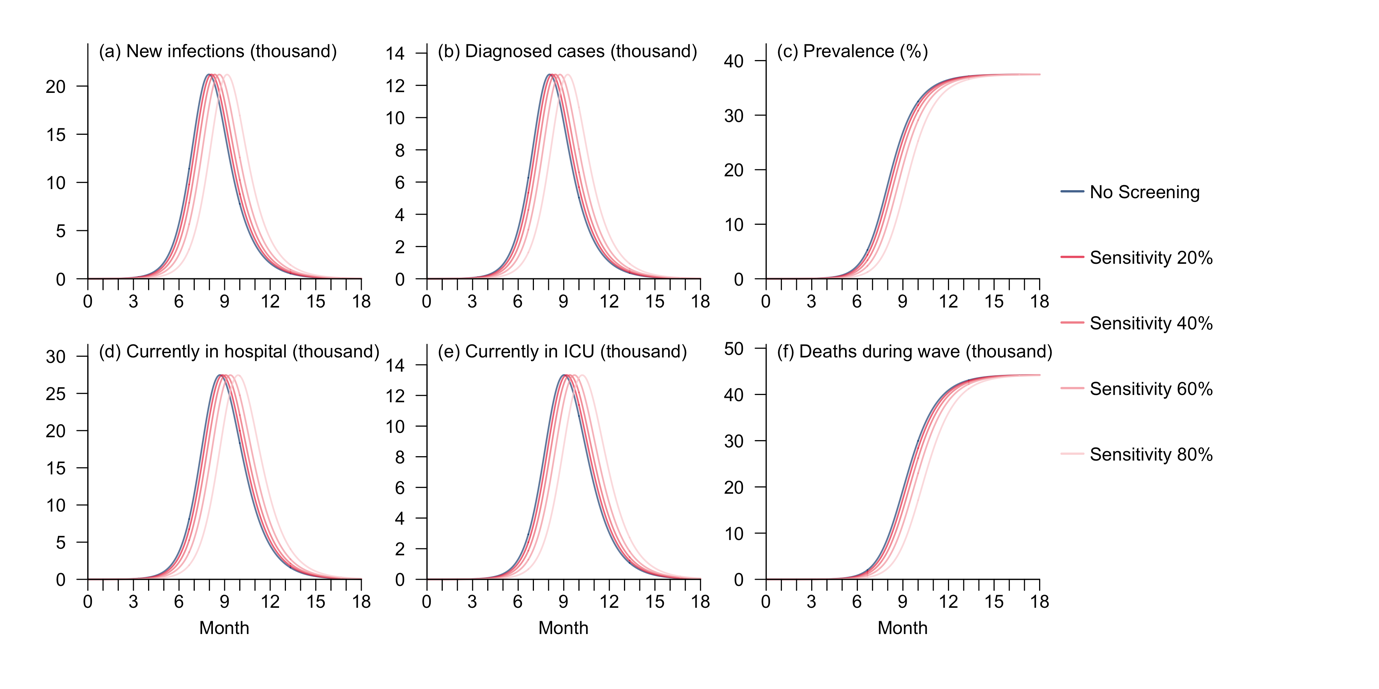

**Figure S6. Disease burden and HCRU for hypothetical mpox outbreaks under scenarios with varying levels of pre-departure testing sensitivity.** The indicators include number of new local infections (a) and diagnosed cases (b) on each day, disease prevalence in the overall local population (c), number of hospitalized cases in normal wards (d) and ICU occupancy (e), as well as cumulative number of deaths (f) over the 18 months following the initial importation event.

**Table S9. Disease burden and HCRU indicators for hypothetical mpox outbreaks under scenarios with varying levels of pre-departure testing sensitivity.** Peak number of new infections and time with maximum number of new infections were calculated for local infections only, while peak number of active hospitalizations (normal ward) and time with maximum hospital occupancy were reported for the overall population.

| **Scenario** | **Peak Infections**  **(n)** | **Time when new infections peaked (month)** | **Peak active hospitalisations**  **(n)** | **Time when hospital occupancy peaked (month)** |
| --- | --- | --- | --- | --- |
| No Screening | 21,208 | 8.0 | 26,643 | 8.7 |
| Sensitivity 20% | 21,205 | 8.1 | 27,469 | 8.9 |
| Sensitivity 40% | 21,200 | 8.3 | 27,465 | 9.1 |
| Sensitivity 60% | 21,196 | 8.6 | 27,461 | 9.4 |
| Sensitivity 80% | 21,195 | 9.2 | 27,456 | 9.9 |

### Sensitivity Analysis: Impacts of diverse NPIs in the extreme scenarios

In the main analysis, we projected the effects of the three NPI strategies on outbreak scales and HCRU in the baseline scenario of moderate disease transmissibility. Nevertheless, the effectiveness of NPIs might also be subject to disease transmission dynamics [7]. The question therefore arises whether these measures would function similarly in extreme scenarios, such as minimal disease transmissibility, which might also be the case for the ongoing mpox spread, as the majority of identified cases outside Africa have been categorised as imported.

To address this question, we performed a sensitivity analysis to project the impacts of the three proposed NPI strategies in the scenario with $R_{s}$ $=4.0$ and $R_{c} =0.1$, while aligning other model parameters with the baseline scenario. Compared to $R_{c}=0.6$, the simulated infection curve for the setting with $R_{c}=0.1$showed a slightly higher increase in the impacts of NPIs on controlling the outbreak, with isolation reducing the maximum number of new local infections by 35.9% and postponing the peak by 2.3 months. Adding screening and contact tracing sequentially would further decrease the peak size by 0.04 and 1.3 percentage points, respectively, and extend the duration between initial importation and the peak of 0.8 and 6.3 months, respectively (Figure S7, Table S10).

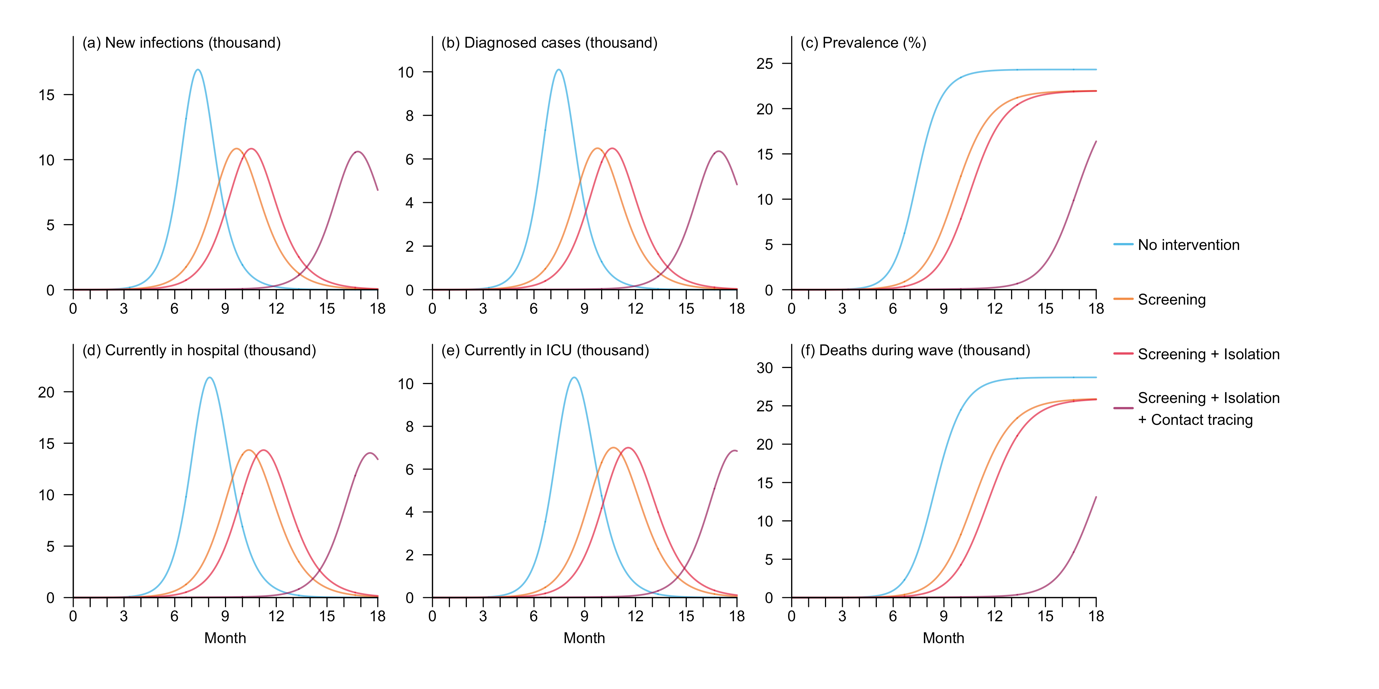

**Figure S7. Disease burden and HCRU for hypothetical mpox outbreaks under scenarios with no intervention or with at least one NPI implemented, assuming extremely low disease transmissibility.** The indicators include number of new local infections (a) and diagnosed cases (b) on each day, disease prevalence in the overall local population (c), number of hospitalized cases in normal wards (d) and ICU occupancy (e), as well as cumulative number of deaths (f) over the 18 months following the initial importation event.

**Table S10. Disease burden and HCRU indicators for hypothetical mpox outbreaks under scenarios with no intervention or with at least one NPI implemented, assuming extremely low community transmissibility.** Peak number of new infections and time with maximum number of new infections were calculated for local infections only, while peak number of active hospitalizations (normal ward) and time with maximum hospital occupancy were reported for the overall population.

| **Scenario** | **Peak Infections**  **(n)** | **Time when new infections peaked (month)** | **Peak active hospitalisations**  **(n)** | **Time when hospital occupancy peaked (month)** |
| --- | --- | --- | --- | --- |
| Baseline | 16,928 | 7.4 | 21,395 | 8.1 |
| Isolation | 10,850 | 9.7 | 14,340 | 10.4 |
| Isolation +  Screening | 10,843 | 10.5 | 14,330 | 11.2 |
| Isolation +  Screening +  Contact tracing | 10,620 | 16.8 | 14,052 | 17.5 |

A similar simulation was conducted to assess the effectiveness of NPIs in the scenario with high non-sexual transmissibility, setting $R_{s}$ = $4.0$ and $R_{c}$ = $1.5$. This meant high overall disease transmission potential in the general population, with spread driven by local infections and non-sexual activities. The simulated epidemic trajectories indicated that these NPIs were slightly less effective compared to the scenario with lower community transmissibility. Specifically, isolation lowered peak number of new local infections by 32.3% and delayed the peak by 1.7 months. While the sequential addition of screening and contact tracing provided only marginal additional benefits relative to isolation in terms of outbreak size, they further postponed the outbreak’s peak by 0.6 and 3.4 months, respectively (Figure S8, Table S11). Nonetheless, it should be noted that based on the most recent mpox case data at the time of this study, such high levels of community transmission appeared to be unlikely.

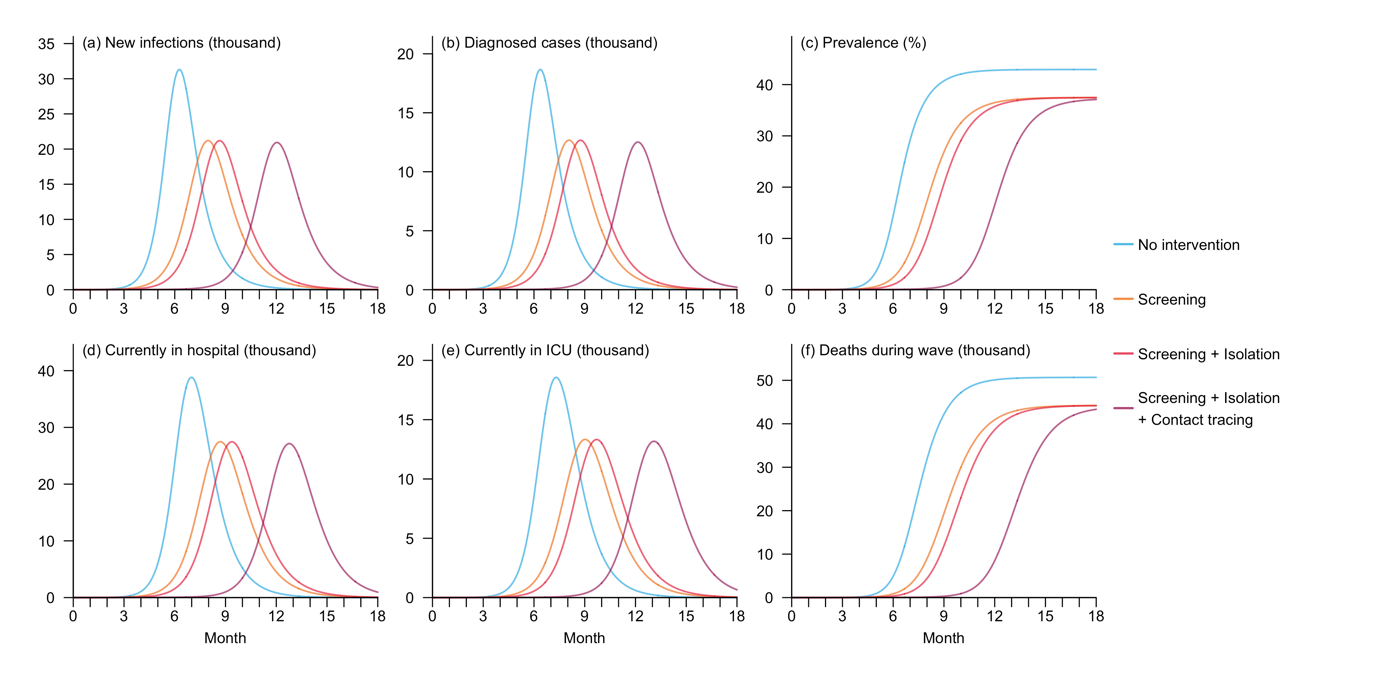

**Figure S8. Disease burden and HCRU for hypothetical mpox outbreaks under scenarios with no intervention or at least one NPI implemented, assuming high community transmissibility.** The indicators include number of new local infections (a) and diagnosed cases (b) on each day, disease prevalence in the overall local population (c), number of hospitalized cases in normal wards (d) and ICU occupancy (e), as well as cumulative number of deaths (f) over the 18 months following the initial importation event.

**Table S11. Disease burden and HCRU indicators for hypothetical mpox outbreaks under scenarios with no intervention or with at least one NPI implemented, assuming high community transmissibility.** Peak number of new infections and time with maximum number of new infections were calculated for local infections only, while peak number of active hospitalizations (normal ward) and time with maximum hospital occupancy were reported for the overall population.

| **Scenario** | **Peak Infections**  **(n)** | **Time when new infections peaked (month)** | **Peak active hospitalisations**  **(n)** | **Time when hospital occupancy peaked (month)** |
| --- | --- | --- | --- | --- |
| Baseline | 31,313 | 6.3 | 38,815 | 7.0 |
| Isolation | 21,208 | 8.0 | 27,473 | 8.7 |
| Isolation +  Screening | 21,196 | 8.6 | 27,461 | 9.4 |
| Isolation +  Screening +  Contact tracing | 20,936 | 12.0 | 27,147 | 12.8 |
